# Optical Genome Mapping: Clinical Validation and Diagnostic Utility for Enhanced Cytogenomic Analysis of Hematological Neoplasms

**DOI:** 10.1101/2022.03.14.22272363

**Authors:** Nikhil Shri Sahajpal, Ashis K Mondal, Tatiana Tvrdik, Jennifer Hauenstein, Huidong Shi, Kristin K. Deeb, Debra Saxe, Alex Hastie, Alka Chaubey, Natasha M. Savage, Vamsi Kota, Ravindra Kolhe

## Abstract

Hematological neoplasms are predominantly defined by chromosomal aberrations that include structural variations (SVs) and copy number variations (CNVs). The current standard-of-care (SOC) genetic testing for the detection of SVs and CNVs relies on a combination of traditional cytogenetic techniques that include karyotyping, fluorescence in situ hybridization (FISH), and chromosomal microarrays (CMA). These techniques are labor-intensive, time and cost-prohibitive, and often do not reveal the genetic complexity of the tumor. Optical genome mapping (OGM) is an emerging technology that can detect all classes of SVs in a single assay. We report the results from our clinical validation (in a CLIA setting) of the OGM technique for hematological neoplasms. The study included 92 sample runs (including replicates) using 69 well-characterized unique samples (59 hematological neoplasms and 10 controls). The technical (QC metrics and first-pass rate) and analytical performance [sensitivity, specificity, accuracy, positive predictive value (PPV), and negative predictive value (NPV)] were evaluated using the clinical samples. The reproducibility was evaluated by performing inter-run, intra-run, and inter-instrument comparisons using six samples run in triplicates. The limit of detection (LoD) for aneuploidy, translocation, interstitial deletion, and duplication was assessed. To confirm the LoD, samples at 12.5%, 10%, and 5% allele fractions (theoretical LoD range) were run in triplicates. The technical performance resulted in a 100% first-pass rate with all samples meeting the minimum QC metrics. The analytical performance showed a sensitivity of 98.7%, specificity of 100%, accuracy of 99.2%, PPV of 100%, and NPV of 98%, which included the detection of 61 aneuploidies, 34 deletions, 28 translocations, 11 duplications/amplifications, 15 insertions/additional material not identified with karyotyping, 12 marker chromosomes, and one each of ring chromosome, inversion and isochromosome. OGM demonstrated robust technical and analytical inter-run, intra-run, and inter-instrument reproducibility. The LoD was determined to be at 5% allele fraction for all the variant classes evaluated in the study. In addition, OGM demonstrated higher resolution to refine breakpoints, identify the additional genomic material, marker, and ring chromosomes. OGM identified several additional SVs, revealing the genomic architecture in these neoplasms that provides an opportunity for better tumor classification, prognostication, risk stratification, and therapy selection. This study is the first CLIA validation report for OGM for genome-wide structural variation detection in hematological neoplasms. Considering the technical and analytical advantages of OGM compared to the current SOC methods used for chromosomal characterization, we highly recommend OGM as a potential first-tier cytogenetic test for the evaluation of hematological neoplasms.

## Introduction

The World Health Organization (WHO) classifies hematological neoplasms into myeloid and lymphoid subtypes, several of which are defined by the presence of specific genetic abnormalities [**1**]. The genetic information provides support to the diagnosis and is important for prognostication, therapy selection, and disease monitoring [**2**]. Genetic diagnostic workup in routine clinical use includes both molecular and cytogenetic analysis to investigate single nucleotide variants (SNVs), small indels, and structural variations (SVs), respectively [**3-7**]. In the past decade, significant progress has been made in the molecular profiling of these tumors with the use of next-generation sequencing (NGS) technology, which has replaced single variant/gene analysis with gene panels and even whole-exome sequencing [**8-9**]. However, since these neoplasms are predominantly defined by chromosomal abnormalities, the cytogenetic analysis remains the primary investigation as recommended by the professional guidelines [National Comprehensive Cancer Network (NCCN) guidelines for myeloproliferative neoplasms (MPN), myelodysplastic syndromes (MDS), acute myeloid leukemia (AML), and lymphomas; National Cancer Institute–Working Group guidelines for chronic lymphocytic leukemia; International Myeloma Working Group (IMWG) guidelines for plasma cell myeloma] [**3-7**,**10**].

The cytogenetic analysis currently relies on a combination of traditional techniques that include karyotyping, fluorescence in situ hybridization (FISH), and chromosomal microarrays (CMA), with each technique having specific utility and limitations. Karyotyping enables genome-wide SV detection but has low-resolution with a maximum banding resolution of ∼5-10 Mb. FISH has higher resolution but is targeted and does not provide genome-wide analysis. CMA has the highest resolution to detect CNVs, but cannot detect balanced SVs (translocations, fusions, and inversions) or determine the location or orientation of copy number gain (or amplification) regions of the genome. As a result, clinically relevant information remains intractable with these current standard-of-care (SOC) techniques. First, though these neoplasms harbor several acquired balanced translocations [**11**,**12**], the detection has remained limited to the recurrent translocations (either cancer-driving gene fusions or overexpression of oncogenes) with targeted FISH panels. Given this limitation, the discovery of new gene fusions has not been possible due to the lack of genic resolution with karyotyping. Second, the location of insertion and orientation of submicroscopic duplications remain beyond the purview of these technologies, which might result in incorrect interpretation of the observed SV. Third, these techniques are not able to resolve complex rearrangements (chromothripsis and chromoplexy) to reveal the chromosomal structure that might add additional clinically relevant information. Considering these limitations with the current cytogenetic analysis, there has been a dire need for a technology that can detect all classes of SVs with a higher resolution in a single assay. The utility of whole-genome sequencing (GS) as a complete solution for genetic analysis was recently tested, but is met with exhaustive bioinformatics and sophisticated instrumentation. These analysis remained limited to the detection of recurrent SVs (selected translocations), and karyotype level CNVs (>5Mb resolution), which does not offer additional benefits over the conventional cytogenetic methods. Furthermore, GS at 50-120x read depth lacks sensitivity to detect certain SNVs, is time and cost-prohibitive, and remains intractable for adoption in routine clinical laboratories [**13**].

In this context, optical genome mapping (OGM) has emerged as a next-generation cytogenomic technology that can detect all classes of SVs at a higher resolution than the SOC techniques. The OGM technique is based on imaging ultra-long DNA (>150 kbp) molecules labeled at a specific 6 bp sequence motif (CTTAAG) that occurs across the entire genome (on average, every 5 kbp). The data generated at approximately 400x genome coverage and the rare variant analysis pipeline ensures the detection of low-level mosaic SVs by comparing single molecules directly with the reference assembly. The accurate and precise location of the labels enables the detection of different classes of SVs including deletions, duplications, and balanced/unbalanced genomic rearrangements (insertions, inversions, and translocations). In addition, a separate coverage-based algorithm detects large CNVs and aneuploidies (similar to CMAs). The variants from the two algorithms are simultaneously visualized in a unique analysis tool for ease of interpretation, variant classification, and reporting (Bionano access 1.6 software). Recently, the technology has gained enormous traction and has been evaluated in several settings, including prenatal [**14**], postnatal [**15**], hematological neoplasms [**16**], and solid tumors [**17**], demonstrating 100% clinical concordance with traditional cytogenetic analysis. However, the literature lacks any clinical validation studies, which are critical for comprehensive evaluation and clinical implementation. In this study, we report the results from our clinical validation (in a CLIA setting) of OGM for hematological neoplasms in comparison to the SOC methods (karyotyping and FISH), which included analyzing technical and analytical performance, reproducibility, and limit of detection for different SV classes. We report robust technical and analytical performance, as well as clinical validity for the utility this platform for hematological malignancies.

## Materials and Methods

### Sample selection

This retrospective validation study included the analysis of 92 samples (including replicates), representing 69 unique and well-characterized samples that were received in our clinical laboratory for cytogenetic analysis with karyotyping and/or FISH testing. These comprised of 59 hematological neoplasms that included adult acute myeloid leukemia (AML) (n=18), chronic lymphocytic leukemia (CLL) (n=15), myelodysplastic syndromes (MDS) (n=12), plasma cell myeloma (PCM) (n=6), lymphoma (n=3), myeloproliferative disorders/myeloproliferative neoplasms (MPD/MPN) (n=3), and chronic myeloid leukemia (CML) (n=2). Additionally, 10 phenotypically “normal” and cytogenetically “negative” samples were also analyzed to evaluate true negative/false-positive rates and calculate performance metrics (**Figure 1a)**. These 69 samples comprised of bone marrow aspirate (BMA) (n=45), peripheral blood (n=16), CD-138 isolated cells (n=5), and lymph node-single cell suspensions (n=3), which were processed for OGM with the technologist blinded to previous SOC results (**Figure 1b)**. Of these 69 samples, 64 were stored at -80°C within 4 days after sample collection (as recommended by the manufacturer), while 6-8 days had elapsed for 3 samples, and 18-20 days had elapsed for 2 samples (**Figure 1c**). The study was performed under IRB A-BIOMEDICAL I (IRB REGISTRATION #00000150), Augusta University. HAC IRB # 611298. Based on the IRB approval, the need for consent was waived; all PHI was removed, and all data were anonymized before accessing the clinical validation study.

**Figure 1.**
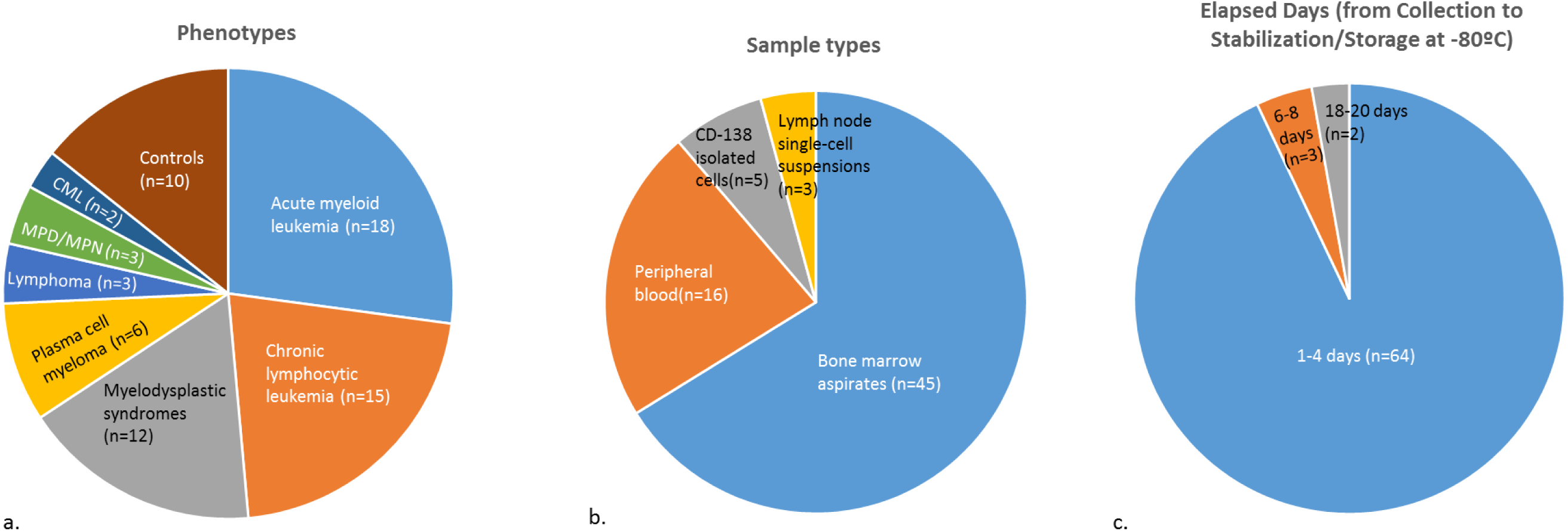
a) The different phenotypes, b) the sample types, and c) storage conditions included in the validation study.

### Optical genome mapping

Ultra-high molecular weight (UHMW) DNA was isolated, labeled, and processed for analysis on the Bionano Genomics Saphyr® platform following the manufacturer’s protocols (Bionano Genomics Inc., San Diego, USA). Briefly, a frozen BMA aliquot (650μl) was thawed and cells were counted using HemoCue (HemoCue Holding AB, Ängelholm, Sweden). Subsequently, a BMA aliquot comprising of approximately million nucleated white blood cells was centrifuged, the cells were digested with Proteinase K (PK), and lysed using Lysis and Binding Buffer (LBB) buffer. DNA was precipitated on a nanobind magnetic disk using isopropanol and washed using buffers (buffer A and B). The UHMW bound DNA was suspended in elution buffer and quantified using Qubit broad range (BR) dsDNA assay kits (ThermoFisher Scientific, San Francisco, USA).

DNA labeling was performed following manufacturer’s protocols (Bionano Genomics Inc., USA) in which 750 ng of purified UHMW DNA was labeled at the sequence specific motif with DL-green fluorophores using Direct Labeling Enzyme 1 (DLE-1) reactions. Following the labeling reaction, the DLE-1 enzyme was digested using PK and the DL-green was removed in two steps using an adsorption membrane in a micro-titer plate. Finally, the DNA backbone was stained blue using DNA stain and quantified using Qubit high sensitivity (HS) dsDNA assay kits. Labeled DNA was loaded onto flow cells of Saphyr chips for optical imaging. The fluorescently labeled DNA molecules were imaged on the Saphyr instrument after the labeled DNA molecules were electrophoretically linearized in the nanochannel arrays. Analytical QC targets were set to achieve >400X effective coverage of the genome, >70% mapping rate, 13-17 label density (labels per 100kbp), and >230 kbp N50 (of molecules >150 kbp).

### OGM variant calling and data analysis

Genome analysis was performed using the rare variant pipeline included in the Bionano Access (v.1.6 or v1.5)/Bionano Solve (v.3.6 or v.3.5) software for all the samples. Briefly, molecules of a given sample dataset were directly aligned to GRCh38, reference human genome assembly, and SVs (insertion, duplications, deletions, inversions, and translocations) were detected based on the differences in the alignment of labels between the sample and the reference assembly. Additionally, a coverage-based algorithm enabled the detection of large CNVs and aneuploidies. SVs and CNVs generated by the rare variant pipeline were then annotated with known canonical gene sets extracted from the reference genome assembly.

For data analysis, the variants were filtered using the following criteria for: 1) the manufacturer’s recommended confidence scores (v.1.6) were applied: insertion: 0, deletion: 0, inversion: 0.01, duplication: -1, translocation: 0, and copy number: 0.99 (low stringency, filter set to 0). 2) The GRCh38 SV mask filter that hides any SVs in difficult to map regions was turned off for analysis. 3) To narrow the number of variants to be analyzed, we filtered out polymorphic variants, i.e. those that appeared in >1% of an internal OGM control database (n>300).

### Performance metric evaluation

Five performance criteria were evaluated viz. positive percentage agreement (PPA), negative percentage agreement (NPA), positive predictive value (PPV), negative predictive value (NPV), and accuracy. For concordance evaluation, the clinically reported variants were compared between OGM and SOC methods, and a variant was considered concordant even if the size or breakpoint was slightly different. Differences in size and breakpoint are anticipated, as the resolution of OGM is higher than SOC methods. However, balanced SVs with centromeric breakpoints, or with a VAF <5% were excluded from the comparison (beyond the detection capabilities of OGM in the current iteration and with 400x coverage).

Additionally, we analyzed each genome for all clinically relevant SVs, and report findings as “concordant” to previous SOC results and “additional findings” that were not reported with SOC methods (**Supplementary file 1)**. Although detailed investigation and confirmation of these additional findings remain beyond the scope of this manuscript, we have provided all clinically reportable SVs in **Supplementary file 1** to assist future discoveries and consider these findings as likely true events, consistent with previous reports [**16**,**17**].

### Analytical comparison between OGM and SOC results

Of the 59 hematological neoplasms, 48 had both karyotyping and FISH, 10 had only FISH, and 1 had only karyotyping in clinical cytogenetic testing (**Figure 2a**). Of the 59 cases, 16 were complex cases (≥4 cytogenetic aberrations) and 43 were simple cases (≤3 cytogenetic aberrations). Overall, the 59 cases harbored a total of 164 aberrations that included 61 aneuploidies, 34 deletions, 28 translocations, 11 duplications/amplifications, 15 insertions/additional material not identified with karyotyping, 12 marker chromosomes, 1 each of ring chromosome, inversion, and isochromosome, which were clinically reported and were included for comparison with OGM results (**Figure 2b**).

**Figure 2.**
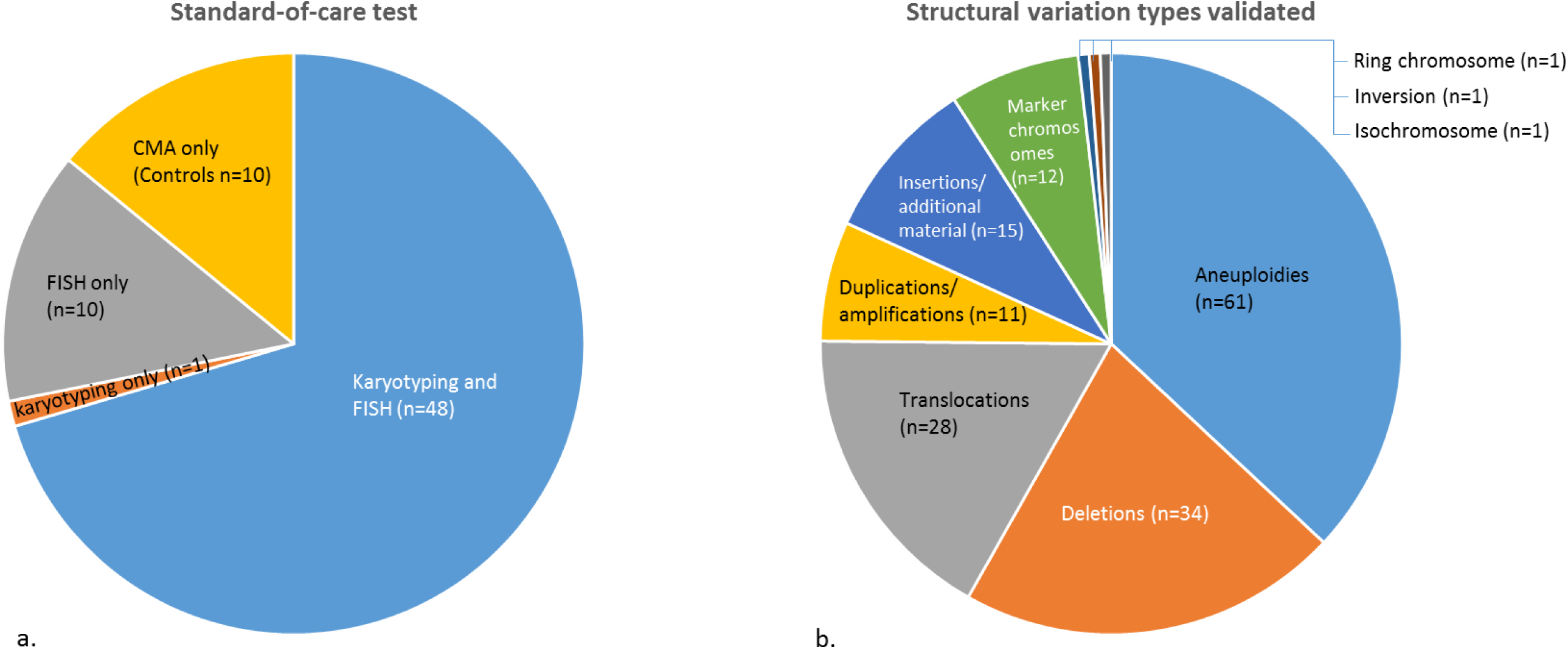
a) The standard-of-care tests compared for concordance and, b) the structural variation types validated in this study.

### Reproducibility studies

The reproducibility was evaluated by performing inter-run, intra-run, and inter-instrument comparisons. To evaluate inter-run and inter-instrument comparison, three samples were run in triplicates on three separate chips and two different instruments. To evaluate intra-run performance, three samples were run in triplicates on the same chip and instrument. The reproducibility was measured for both technical (QC metrics) and analytical (clinically reported variant) performance.

### Limit of detection studies

The limit of detection (LoD) of the OGM platform at the set parameters (400x genome coverage, rare variant pipeline, and variant confidence threshold) were assessed for aneuploidy, translocation, interstitial deletion, and duplication. The LoD studies were performed for each variant class by diluting a sample with known variant and allele fraction (reported by SOC method) and diluting with wild type DNA to provide the following allele fractions 25%, 16.6%, 12.5%, 10%, and 5%. The samples at 12.5%, 10%, and 5% allele fractions (theoretical LoD range) were run in triplicates to confirm the LoD.

## Results

### Technical Performance: OGM quality control metrics and variant filtering

A typical run for OGM included the processing of eight samples in a batch for DNA isolation, and 12 samples for labeling, while three samples were loaded into one nanochannel array chip and two chips onto the Saphyr instrument at a given time. All 69 samples passed the quality control metrics and the 59 hematological neoplasm samples achieved an average N50 (>150 kb) of 303 kb (±35), map rate of 87.5% (±7.5), label density of 15.8/100 kb (±1.0), and average coverage of 391x (±89) (**Supplementary file 2**). In total, 86306 SVs were identified in the 59 samples, with an average of ∼1462 SVs per sample. Of all the identified SVs, a total of 44226 SVs remained after the filtration criteria were applied (94.9% variants filtered out), with an average of ∼71 SVs per sample that were further interrogated **(Supplementary file 3)**.

### Analytical performance: Comparison with SOC results

The 59 hematological neoplasms consisted of simple (n=43) and complex cases (n=16), which harbored all classes of SVs (61 aneuploidies, 34 deletions, 28 translocations, 11 duplications/amplifications, 15 insertions/additional material not identified with karyotyping, 12 marker chromosomes, 1 each of ring chromosome, inversion and isochromosome), and were representative of the variety of SVs observed in any clinical laboratory. OGM was concordant in identifying 162/164 variants, which were reported with current SOC methods. The two variants that were not detected with OGM included mosaic loss of chromosome Y and an insertion, both detected at <10% allele fraction with karyotyping, in two complex cases of CLL. The two variants were detected in a low fraction of cells (4/20, and 3/20) suggesting a varying proportion of abnormalities in the proliferative clones vs in the extracted bulk DNA.

These different classes of SVs also included the well-known aberrations in hematological neoplasms. As such, the 61 aneuploidies included 33 monosomies, 21 trisomies and 8 loss of sex chromosomes, of which the most common were monosomies 5 (n=5), 7 (n=5), 13 (n=5), trisomies 8 (n=3), 9 (n=3), 11 (n=3), 21 (n=3), and loss of Y (n=7) chromosome. OGM detected 60/61 aneuploidies, while one mosaic loss of chromosome Y (in a complex case of CLL) was not detected with OGM. Of these 60 aneuploidies, the software correctly called 58 aneuploidies, while two were manually inferred (and detected at a low confidence threshold) from the visualization in the whole genome CNV viewer. These two aneuploidies, +12, and -13, were reported at the LoD range with ≤ 5% allele fraction with karyotyping (detected in a low fraction of cells (4/20, and 3/20, respectively).

The 34 deletions included in the study ranged from 905 kb up to the loss of an entire chromosome, which were uniquely called by both the CNV and SV algorithm in the OGM data. OGM concordantly characterized the 29/29 translocations that included balanced, unbalanced, and three-way translocations, including those that lead to common gene fusions viz. *RUNX1::RUNX1T1* (n=3), *KMT2A::ELL* (n=1), *BCR::ABL1* (n=1) and *FGFR1::BCR* (n=1) (**Figure 3a-d**). Further, OGM detected the duplications and was able to identify the 14/15 additional material/insertions, revealing the identity of the 12 markers and 1 ring chromosome that remained uncharacterized by karyotyping (**Figure 4a-d**).

**Figure 3.**
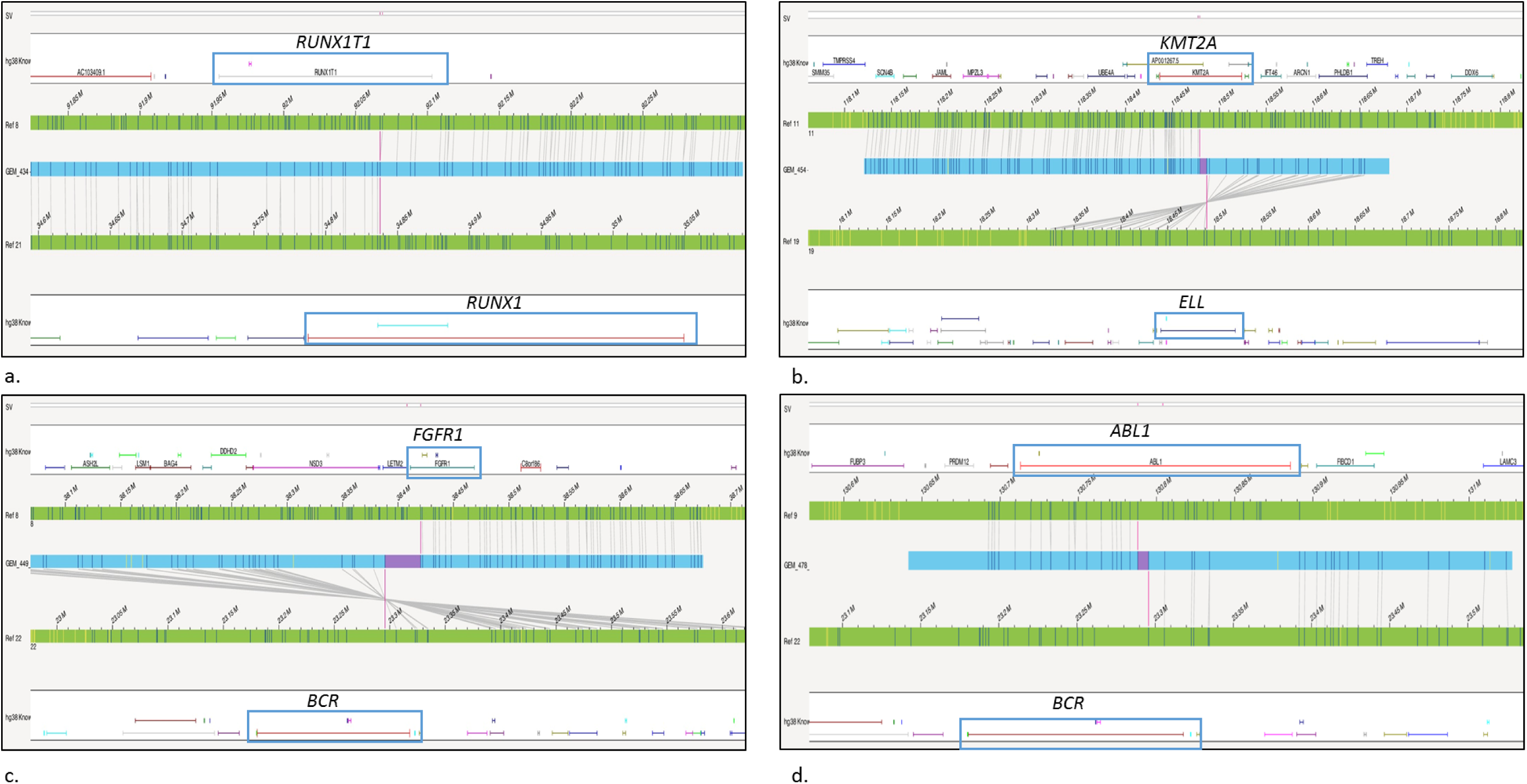
Representative examples of common gene fusion detected with optical genome mapping a) *RUNX1::RUNX1T1*, b) *KMT2A::ELL*, c) *FGFR1::BCR, and d) BCR::ABL1*.

### OGM in simple cases: 100% concordance and higher resolution

Of the 43 cases classified as simple, 35 cases had at least one clinically reported genetic aberration, while eight cases were negative with both karyotyping and FISH testing. In these 35 cases, OGM detected all the previously reported variants and even corrected putative false-positive results in two cases:

> Case 1: In a case of multiple myeloma with a negative karyotype (performed on BMA), the FISH testing (performed on CD-138 cells) reported a gain of one to two copies of 1q *CKS1B* (43 of 50 cells), monosomy 13 (47 of 50 cells), a gain/rearrangement of one copy of 4p *FGFR3* (23 of 50 cells). OGM (performed on CD-138 cells) confirmed 1q21.1qter(144092961_248943333)x3, (13)x1, but did not detect a copy number gain or rearrangement of the *FGFR3* gene. However, OGM detected a *der(4)t(4;5)(p16*.*3;p13*.*3)(1910123;32591078)*, with the breakpoint at 4p16.3 that overlaps the region of *FGFR3* dual fusion probes used for detecting *FGFR3* gain/rearrangement. However, the breakpoint of the translocation did not disrupt the *FGFR3* gene **(Figure 5a-b**).

**Figure 5.**
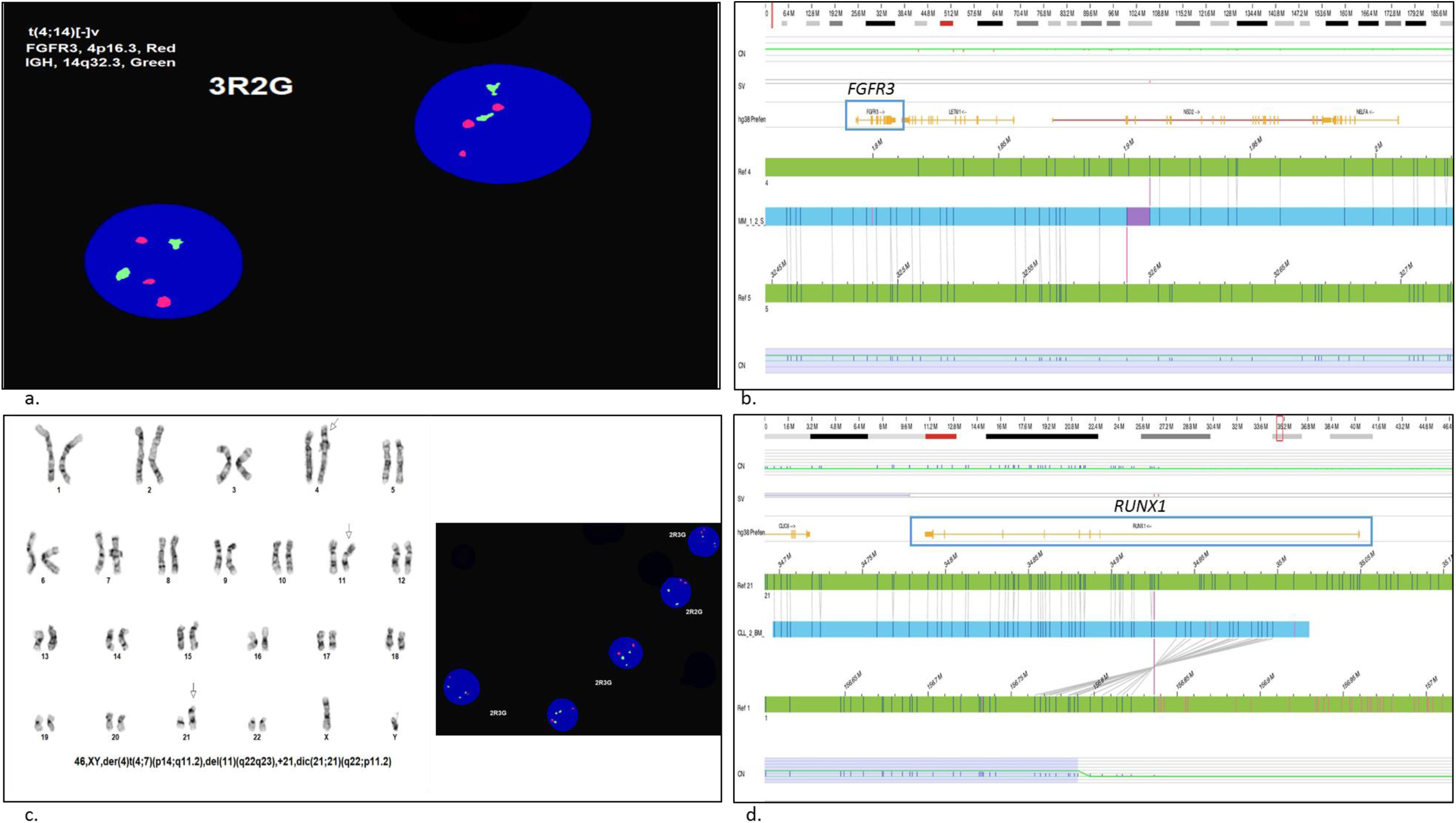
Optical genome mapping corrected false-positive results in two cases. a) In a case of multiple myeloma, the FISH testing shows a gain of one copy of 4p *FGFR3* (23 of 50 cells). b) OGM revealed that the gain observed in FISH was because of the t(4;5)(p16.3;p13.3)(1910123;32591078), with the breakpoint at 4p16.3 that overlaps the region of FISH dual fusion probe used for detecting *FGFR3* gain. The breakpoint of the translocation did not disrupt the *FGFR3* gene. c) In a case of CLL, Karyotyping detected dic(21;21)(q22;p11.2)[12/20], and interphase FISH with *RUNX1* probe showed gain of *RUNX1*. d) OGM results indicate that the dic (21;21) is most likely a der(21)ins(21;1)(q22.12;q?21.1q23.1)inv dup(q?21.1q23.1) with a copy number gain at the breakpoint at chr 1 [1q21.1q23.1(144173.283_156790.211)x4].

> Case 2: In a case of CLL, karyotyping detected 46,XY,der(4)t(4;7)(p14;q11.2),del(11)(q22q23), *dic(21;21)(q22;p11*.*2)[12]/*46,XY[8], and FISH confirmed the deletion at 11q (ATM). OGM was concordant in detecting der(4)t(4;7)(p15.1;q21.11)(29865036;83430032), and 11q22.3q23.3(104626126_117138719)x1. There was a discrepancy in the karyotyping and OGM results for the dic(21;21)(q22;p11.2) anomaly. OGM results indicate that the dic (21;21) is most likely a *der(21)ins(21;1)(q22*.*12;q?21*.*1q23*.*1)inv dup(q?21*.*1q23*.*1)*, with a copy number gain at the breakpoint at chr 1 [1q21.1q23.1(144173.283_156790.211)x4]. The karyotyping was reviewed again in the light of OGM and FISH data that indicated a gain of *RUNX1*, and the der(21)ins(21;1) observed with OGM seems to be the most likely explanation of the observed karyotype. Notably, the OGM data is not able to account for the heterochromatin, whereas the karyotyping is unable to detect the cryptic rearrangement. Given the band resolution of karyotyping (>5Mbp), the OGM results most likely refine and better characterize the abnormality involving chr 21, which disrupts the *RUNX1* gene at intron 1, likely the reason for splitting the *RUNX1* FISH probe in two halves **(Figure 5c-d**).

Overall, OGM was found to be 100% (56/56) concordant in detecting the previously reported variants with the current SOC methods, which included 14 aneuploidies, 13 deletions, 19 translocations, 8 duplications/amplifications, and 1 each of isochromosome and inversion in these cases.

### Additional findings with OGM in simple cases

Of the 43 cases classified as simple, 35 cases had both karyotyping and FISH results. While these 35 cases were classified as simples (<3 aberrations) with cytogenetic analysis, seven of these cases harbored ≥4 aberrations with OGM. For example, in the case of CLL, karyotyping revealed a negative profile with only five metaphases analyzed because of poor sample quality, while FISH reported the loss of 11q and 13q. OGM detected the two aberrations with higher resolution (refined the breakpoints) and detected five additional SVs including chromoanagenesis event at 8q. Of the seven cases that had only FISH results, five were found to have ≥4 aberrations with OGM. Overall, OGM detected many additional SVs including 68 additional translocations, with 38 potential gene fusions in these 43 cases.

In the nine negative cases based on karyotyping and FISH testing, OGM detected cytogenetic aberrations in three cases, i.e. 30%. In case 1 (MDS), OGM detected a t(2;22)(q14.1;q13.33)(113658791;50631611), in case 2 (AML), OGM detected a duplications that disrupted the *MYH9* gene (duplication overlapping exons 6-41). Although, overexpression of *MYH9* gene has been associated with poor prognosis in AML [**18**], it would be interesting to follow up this patient and correlate the prognosis with additional functional analysis (beyond the scope of the present study). In case 3 (MDS), an abnormality fus(17;17)(p11.2q22)(17374798;57608225) disrupting the *MSI2* gene at intron 8 was detected. The *MSI2* expression is required for maintaining activated MDS stem cells and high gene expression is associated with faster progression to AML and is associated with poor prognosis [**19**]. Notably, two novel *MSI2* gene fusions have been very recently reported with OGM in two cases of CML [**16**], and while the functional implications were not assessed, it certainly seems that *MSI2* gene rearrangements are more common than reported in the literature primarily because of the low resolution of SOC methods. It is expected that with the use of OGM, these rearrangements will be better documented and open the avenue for functional characterization that might lead to better prognostication.

### OGM in complex cases: concordance and additional finding

The 16 complex cases harbored 108 genetic aberrations reported with current SOC methods. Of these, 15 cases were found to be in full concordance with previous findings. In these 15 cases, 2 variants required a lower aneuploidy confidence threshold setting for their detection (a loss of chromosomes 13 and gain of chromosome 12). The two variants were detected in a low fraction of cells (4/20, and 3/20) suggesting a varying proportion of abnormalities in the proliferative clones vs in the extracted bulk DNA. Apart from the concordance, OGM was able to reveal the identity of 12 marker chromosomes and one ring chromosome (**Figure 4a-d**). Of these, 11 marker chromosomes and the ring chromosome were found to have complex rearrangements (chromoanagenesis events), while one marker chromosome had resulted from a single unbalanced translocation t(5;17)(q11.2;p11.2)(53194444;22623858). Further, OGM identified eight additional chromoanagenesis events in 4 cases.

OGM refined the breakpoints of translocations detected with karyotyping, and identified putative novel gene fusion *SUGCT::PARP4* in a case of CLL, disrupting the *PARP4* gene at intron 27. In addition, OGM identified several additional SVs, including 145 translocations and 67 novel gene fusions (**Supplementary file 1**). In one notable case, OGM detected two novel fusions *CHD7::FLT3, RAB2A::FLT3*, disrupting both alleles of *FLT3* at intron 3 and intron 4, respectively (**Figure 6a and b**). While confirming the additional findings were beyond the scope of the present study, the results are in alignment with the recent report investigating hematological neoplasms [**16**].

**Figure 6.**
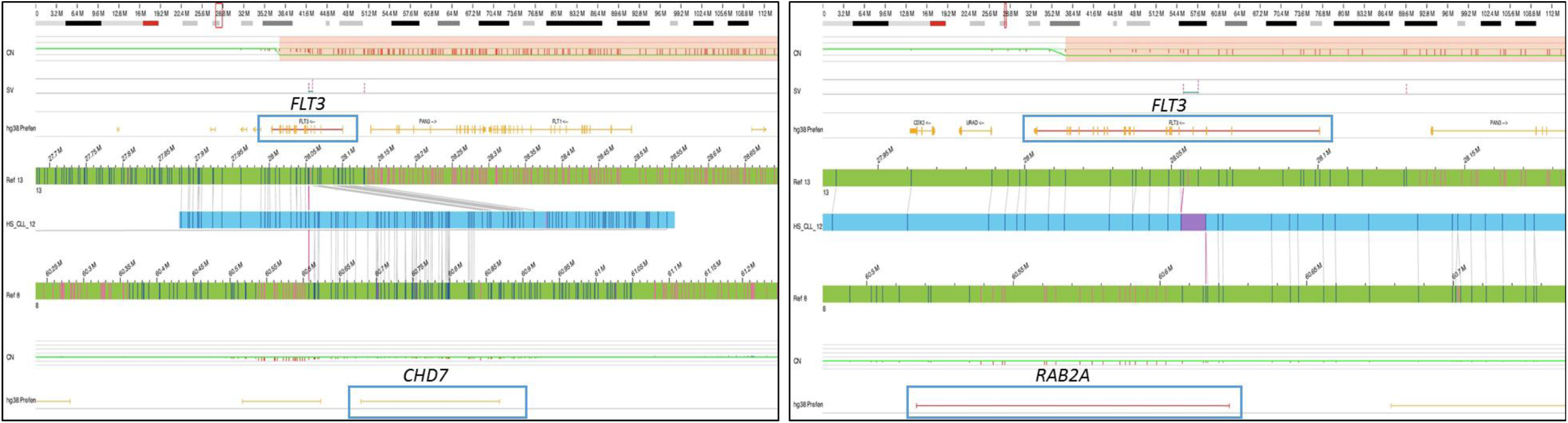
In a case of CLL, OGM detected two novel fusions disrupting both alleles of the *FLT3* gene a) *CHD7::FLT3* disrupting *FLT3* at intron 3 and b) *RAB2A::FLT3* disrupting *FLT3* at intron 4.

### Performance metric evaluation

The performance metrics were calculated for 59 cases that included 59 cases of hematological neoplasms and 10 negative cases of phenotypically normal controls. The 164 known aberrations detected with karyotyping and FISH in the hematological neoplasm cases were used to calculate the true positive rate. OGM was concordant in detecting 162/164 aberrations. Of these 162 variants, OGM corrected two calls made by SOC methods, which were confirmed by additional reviews and were considered true positives. As OGM has higher sensitivity/resolution and has been shown to detect additional variants compared to SOC methods, only the control samples were used for FP and TN calculations, where polymorphic variants were excluded and only oncogenic SVs were considered for calculation. No oncogenic SV was detected with OGM in these cases, consistent with CMA results. From the 166 variants analyzed in this study, OGM was found to have a sensitivity of 98.7%, specificity of 100%, positive predictive value (PPV) of 100%, negative predictive value (NPV) of 98%, and accuracy of 99.2%.

### Reproducibility studies

As part of this study, six samples were in triplicate to evaluate inter-run, intra-run, and inter-instrument reproducibility. All 18 replicates passed the technical QC metric of >400X effective coverage of the genome, >70% mapping rate, 13-17 label density (labels per 100kbp), and >230 kbp N50 (of molecules >150 kbp) (**Supplementary file 4**). These six samples included six translocations (including a three-way translocation), two aneuploidies (one each of monosomy and trisomy), and two interstitial deletions reported with SOC methods. OGM detected these respectively reported variants in each replicate, demonstrating a 100% inter-run, intra-run, and inter-instrument technical and analytical reproducibility (**Supplementary file 5**). In addition, the additional variants detected with OGM in these cases were also investigated and were found to be reproducible across the replicates, demonstrating high overall reproducibility.

### Limit of detection studies

The limit of detection studies were carried out to ascertain the lower LoD of the platform. The LoD was evaluated for the following variant classes: aneuploidy, translocation, interstitial deletion, and duplication. Of these, the translocation, interstitial deletion, and duplication were detected consistently from 25% to 5% allele fraction. The aneuploidy was called by the software till 12.5%, but a lower aneuploidy threshold setting was used to call the aneuploidy at 10% and 5% allele fraction. Overall, these variants were detected in the triplicates at 12.5%, 10%, and 5% allele fraction, demonstrating an LoD of 5% allele fraction at 400x genome coverage with the rare variant pipeline (**Figure 7a-t**).

**Figure 7.**
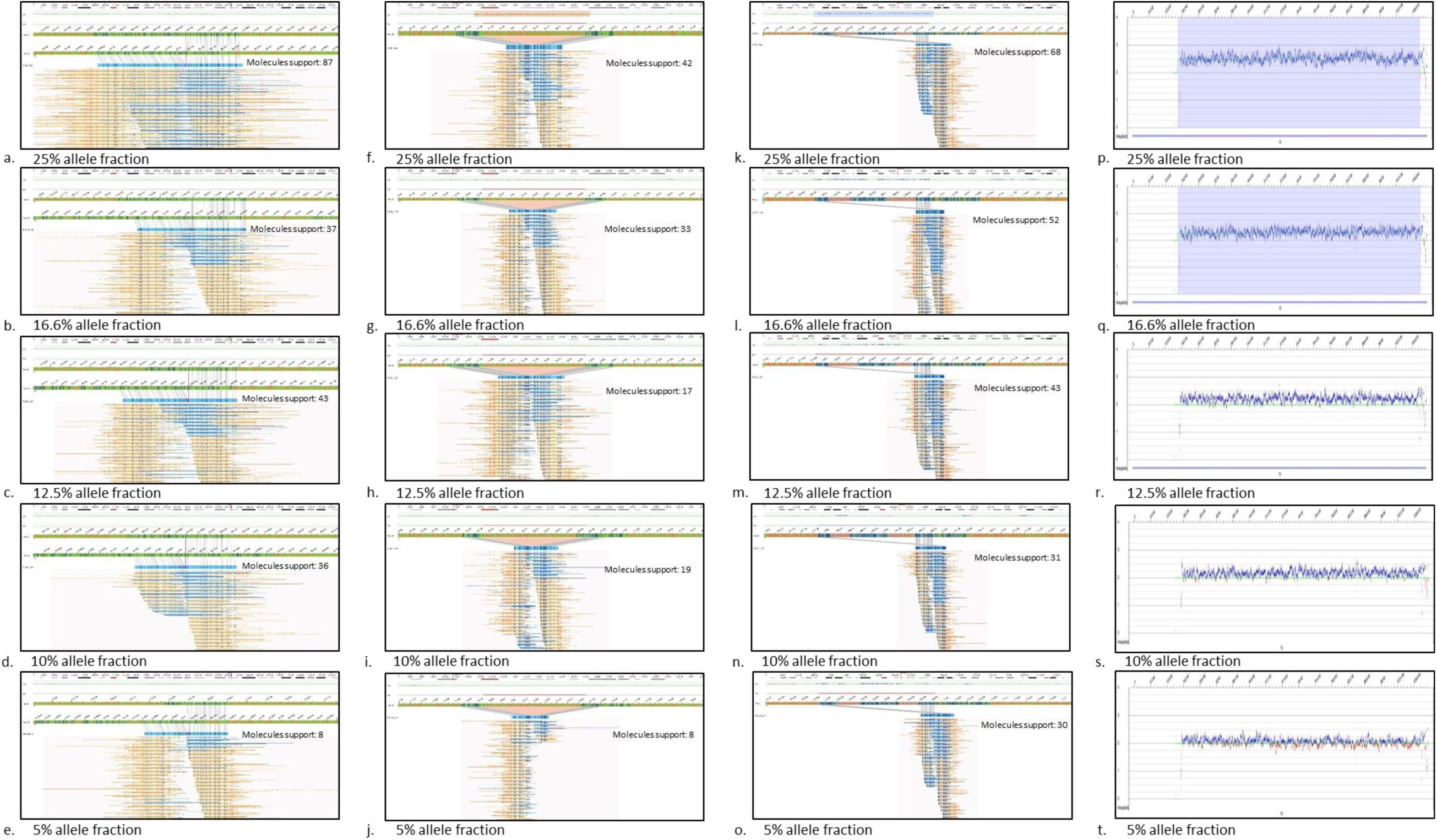
The limit of detection of optical genome mapping for aneuploidy, translocation, interstitial deletion, and duplication was assessed from 25% to 5% allele fraction and was found to be 5% allele fraction for all variant classes. a-e) translocation: t(4;15)(122684318; 25010247) f-j) Interstitial deletion: 17p12p11.2(15752128_16669511)x1, k-o) Interstitial duplication: 2q12.3q13(108513707_110504318)x3, p-t) aneuploidy: trisomy 13.

## Discussion

The cytogenetic analysis of hematological neoplasms is of critical importance for accurate diagnosis, classification, prognostication, therapy selection, and disease monitoring. The professional society guidelines recommend the use of karyotyping and FISH for MDS, MPN, AML, and CLL, while FISH is the primary genetic test for MM [**3-7**]. However, these approaches are not able to detect novel gene fusions, assess if the structural variants are disrupting a gene, or detect submicroscopic aberrations that are not known/included in FISH panels. These limitations are only partially addressed with CMAs, which although facilitate the detection of CNVs at a higher resolution compared to karyotyping, cannot detect balanced rearrangements. To overcome the limitations with current SOC methods and considering the importance of consolidated, high-resolution chromosomal characterization in hematological malignancies, there has been a significant interest to validate OGM technology, as it detects all classes of SVs in a single assay. This study reports and discusses the strategy and results of clinical validation of OGM as per CLIA requirements for a laboratory-developed test (LDT).

### The technical performance of OGM

The sample processing for OGM is simple and standardized for the different sample types analyzed in this study (BMA, peripheral blood, CD-138 cells, and lymphoma-single cell suspensions). These samples are representative of the different sample types processed in any clinical laboratory analyzing hematological neoplasms. The manufacturer’s protocol for blood and BMA can be adapted with a single additional step for CD-138 and lymphoma-single cell suspensions. The feasibility of using these sample types for genome-wide analysis using OGM offers several advantages over the traditional methods, where the testing is currently limited to FISH or CMA (or CD-138 cells, and lymphoma-single cell suspensions), as these cannot be analyzed with karyotyping. Typically, 16 samples were isolated in a single day in two batches of eight samples, and 12 samples were labeled the following day in a single batch by a single technologist. Overall, 12 samples were ready for imaging, with a total hands-on time of approximately 5 hours. Batches of three samples were loaded on individual Saphyr chips and imaged on the instrument in a single run, with ∼1500 gigabasepairs of DNA collected for each sample enabling an average depth of coverage of >400x, with the entire OGM workflow from sample to SV calling accomplished in 4-5 days.

All 92 sample runs passed the pre-analytical and analytical QC metrics, with a first pass success rate of 100%, which highlights the ease of use and demonstrates the feasibility of processing these samples for cytogenetic analysis in a routine clinical laboratory. Further, the samples analyzed in the study were stored at 4°C from two to 20 days before freezing at -80°C, to replicate the real-clinical scenario where delays are caused due to several unforeseeable factors. Remarkably, the QC metrics passed for all samples, except that the one-sample stored until 20 days required a longer time for data collection over two flow cells to reach 400X coverage. Overall, these results demonstrate the robustness of the OGM platform to analyze different sample types including compromised samples for genome-wide SV analysis.

### Analytical performance of OGM compared to SOC methods (karyotyping and FISH)

OGM demonstrated excellent analytical performance, with 98.7% sensitivity, 100% specificity, PPV, 98% NPV, and 99.2% accuracy, detecting 162/164 genetic aberrations in the 59 hematological neoplasm samples.

The two variants, mosaic loss of chromosomes Y and insertion that could not be detected with OGM were detected at 3/29 and 3/20 cells with karyotyping, respectively, which could be explained by a selective bias in the growth of a single clone in culture (note: OGM is performed without culture). Further, the LoD for OGM in this study is 5%, right at the level estimated by the karyotype. It is noteworthy that the 164 aberrations tested in this study ranged from 0.75% to 50% allele fraction, and OGM detected the genetic aberration (loss of 13q with the FISH probe for *DLEU* gene) even at 0.75% allele fraction.

We included markers, additional material, and ring chromosome in the concordance calculations to evaluate if OGM can discern these complex events and reveal important details of genomic rearrangement in these samples. OGM was able to reveal the identity of 15 additional material, 12 marker chromosomes, and the ring chromosome that were refractory to characterization with SOC methods. The marker chromosomes are believed to result from chromoanagenesis and are associated with poor prognosis [**20**]. However, one of the marker chromosomes resulted from a single translocation, while the remaining 11 resulted from chromothriptic events. The case with the marker chromosome that resulted from single rearrangement had a complex karyotype with 6 aberrations {45,XX,-7[9]/44,XX,-5,del(7)(q11.2),-17,-18,+mar[3]/46,XX[8]}, while OGM resolved these 6 aberrations into 3 events. Thus, the complex karyotype, in this case, was rather a simple profile with the marker chromosome that did not result from chromothripsis. Additionally, as no technology was able to completely reveal the identity of marker chromosomes, only the “detection” of marker chromosomes was associated with prognosis [**20**,**21**]. As we gather more data with OGM, we expect to uncover a new and more precise correlation between different marker chromosomes and patient outcomes.

Notably, we also validated the software (version 1.6), confidence threshold for different variant types, and the filtering criteria, which led to a true positive rate of 97.5% (160/164), and a true negative rate of 100%. Apart from the two missed variants (loss of Y and insertion), two aneuploidy calls (chromosomes +12 and -13) were called only after lowering the aneuploidy confidence threshold (both were found at low clonal fractions by karyotype). Although the software called several aneuploidies at similar allele fractions in other cases, we also observed in the LoD analysis that the aneuploidy was called at 10% and 5% allele fractions only after reducing the confidence threshold. It is important to highlight that the analyst (blinded to SOC results) reviewing these cases called these aneuploidies during the manual review, as the evidence for the aneuploidies was apparent using the CNV visualization tool. With these observations, we recommend the use of the default confidence thresholds but also including a manual review of each case for aneuploidies using the whole genome CNV view in the software, which is very similar to the whole genome CNV visualization with CMAs and is a common practice by analysts and directors reporting cases on CMAs. The default confidence thresholds and the manual reviews enabled 98.7% TP and 100% TN rates in this study.

Apart from the high concordance for oncogenic abnormalities, the 0% false-positive rate with the evaluation of 10 phenotypically normal and cytogenetically negative controls is reassuring, as this is an important parameter in the evaluation of any platform. Overall, OGM demonstrated an excellent analytical performance evaluated across all classes of SVs detected in this wide variety of clinical samples.

### Reproducibility and limit of detection

The OGM platform showed high reproducibility at the level of inter-run, intra-run, and inter-instrument, with all triplicates passing the QC parameters and all reported variants detected in respective samples, demonstrating the robustness of the assay. The reproducibility was also confirmed with the LoD studies, where the four variants were consistently detected in the triplicates at 12.5%, 10%, and 5% allele fractions. This is the first study to formally assess the LoD, which was found to be at least 5% allele fraction for aneuploidy, translocation, interstitial duplication, and deletions, at ∼ 400x genome coverage with the rare variant pipeline.

### Higher resolution of OGM: advantages over SOC methods

In simple cases, defined as having ≤3 aberrations by karyotyping/FISH, OGM was 100% concordant in identifying the 56 aberrations detected with SOC methods. In addition, OGM detected several additional aberrations with 12 cases with >4 aberrations and six cases with >7 aberrations). Currently, the definition of a “simple” case is defined as having ≤3 aberrations and the karyotype profile is used for classification/risk stratification according to the NCCN and WHO guidelines. Since OGM reveals additional genomic complexity in these neoplasms, a re-definition of simple vs. complex profiles needs to be established based on the emerging data. The accurate assessment of cytogenetic aberrations with OGM will help to better classify these neoplasms into correct risk categories.

Importantly, OGM refined breakpoints for several SVs and corrected two calls (one detected with karyotype and one with FISH) altering the clinical significance for those SVs. A 4p*FGFR3* gain detected by FISH using double-fusion probes was found to be a false call, as OGM showed that the multiple signals with FISH resulted from a translocation break in between the two parts of FISH double-fusion probe used for detecting *FGFR3* gain or translocation. The breakpoint of the translocation did not disrupt the *FGFR3* gene. In another case, the dic(21;21)(q22;p11.2) abnormality was refined as a ***der(21)ins(21;1)(q22*.*12;q?21*.*1q23*.*1)inv dup(q?21*.*1q23*.*1)*** with a copy number gain at the breakpoint at 1q42.12q42.2 (**Figure 5d**), demonstrating the power of OGM in accurately characterizing variants detected by low resolution karyotyping. This translocation disrupted the *RUNX1* gene at **intron 1**, which to our knowledge has not been reported in CLL, and *RUNX1* gene mutation has been associated with distinct clinicopathologic features and poor prognosis in AML [**22**].

The higher resolution of OGM and advantages over SOC methods can be clearly illustrated by the analysis of 13q14 deletions in CLL cases. The 13q14 deletions are the most common cytogenetic aberrations observed in CLL (>50% of cases) [**23**] and are associated with a good prognosis if detected as a sole aberration by FISH [**24**]. However, the clinical course in these cases has remained heterogeneous because of varying deletion sizes, deletion types (mono or bi-allelic), and additional aberrations in the tumor [**24**,**25**]. Of the 15 CLL cases analyzed in this study, 10 had the 13q14 deletion detected by FISH testing. OGM detected all 10 deletions, identifying multiple clones down to 0.7% allele fraction and better characterized the deletions, six were small deletions overlapping miR15a/16-1 only and four were larger deletions that encompassed miR15a/16-1 and the *RB1* gene (**Figure 8**). The deletion of the *RB1* gene along with miR15a/16-1 has been associated with poor prognosis, while the deletion of miR15a/16-1 deletion alone reflects a good prognosis in CLL cases [**24**,**25**]. One of these cases revealed a complex cytogenetic profile with OGM. This further exemplifies the utility of OGM compared to SOC methods, as it enables the genome-wide detection of all classes of SVs, which can better assist the physician in correlating the genetic profile with disease progression/response to therapy compared to a targeted approach.

**Figure 8.**
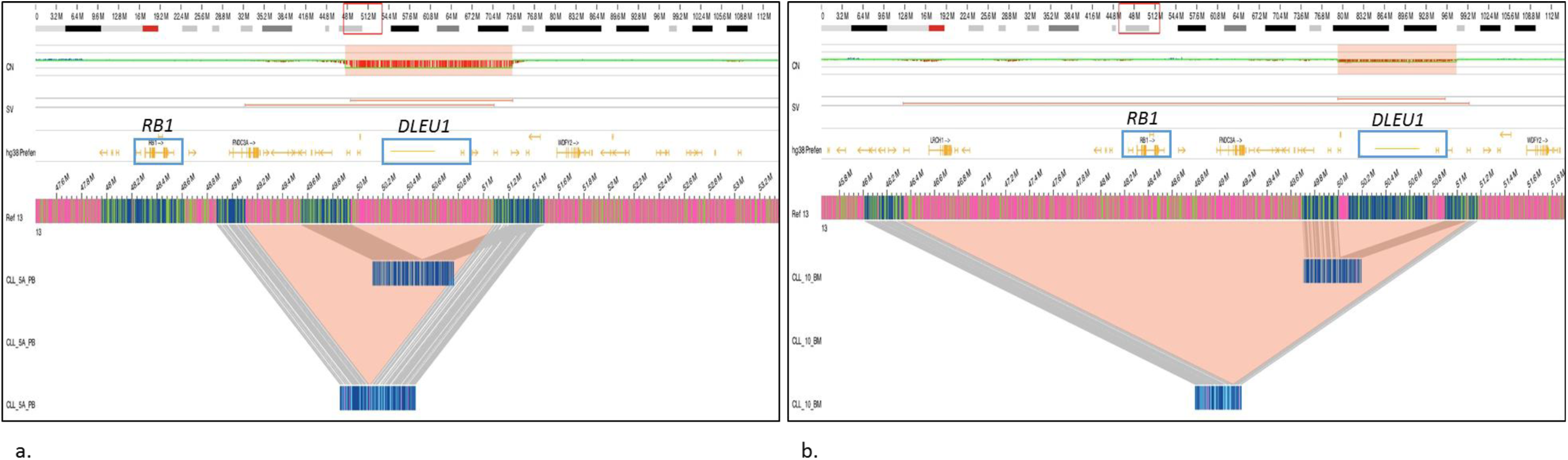
Optical genome mapping can distinguish the different 13q14 deletions that enables accurate prognostication in CLL cases as compared to karyotyping and FISH testing. a) OGM detects two clones with both deleting the 13q14 region overlapping the *DLEU1* gene (miR15a/16-1). b) OGM detects two clones with deletion of 13q14 region, with one clone overlapping the *DLEU1* gene (miR15a/16-1) and second clone encompassing both *DLEU1* gene (miR15a/16-1) and the *RB1* gene.

Another significant advantage of OGM is the ability to detect balanced and unbalanced events in one assay. OGM refined the breakpoints of all the translocations detected with karyotyping, and identified a novel gene fusion *SUGCT::PARP4* in a case of CLL, disrupting the *PARP4* gene at intron 27 that has not been reported before in the literature. OGM detected 202 additional translocations and 104 novel putative gene fusions, which included two novel fusions *CHD7::FLT3* and *RAB2A::FLT3*, disrupting both alleles of *FLT3* at intron 3 and intron 4, respectively (**Figure 6**). These fusions have not been reported in the literature, while other novel *FLT3* gene fusions have been reported in CML, MPN, and chronic myelomonocytic leukemia [**26-28**]. These tumors have shown enhanced sensitivity to *FLT3* inhibitors in *in-vitro* studies, and thus, might have a potential role in therapy decision-making [**26**]. Traditionally, the gene-fusions have been characterized using FISH testing (in some cases with PCR) by identifying one or both fusion partner genes. However, this approach does not facilitate the discovery of novel fusions and is limited to the detection of well-known, recurrent gene fusions. It may also result in false-positive calls, as the breakpoints may not overlap the gene but still lie within the FISH probe region. Short-read RNA sequencing has also been used but remains limited to targeted panels. Recently, GS has been reported to detect cytogenetic aberrations in myeloid neoplasms. While the results are encouraging, the study only assessed recurrent SVs and CNVs smaller than 5Mb. It excluded smaller CNVs, additional fusions, and complex rearrangements, and carries a high cost, and requires exhaustive bioinformatics [**13**]. Further, the sequencing technologies do not have the ability to sequence repeat elements that are longer than read lengths and are intractable to SV in detection in these regions even with long-read sequencing technologies, as shown by recent benchmark studies [**29**,**30**]. Thus, this unique unparalleled ability of OGM to detect all classes of SVs including balanced and unbalanced events in a single assay demonstrates its clinical utility for chromosomal characterization.

## Conclusion

This study is the first CLIA validation report for optical genome mapping for genome-wide structural variation detection in hematological neoplasms. The study shows a 98.7% sensitivity and 100% specificity for detecting SVs previously reported with a combination of SOC methods. The increased clinical utility of OGM in hematological malignancies has been established by multiple reports where 100% concordance was reported with multiple SOC methods. OGM refined the breakpoints in several of these previously reported aberrations and modified the clinical relevance of these variants. OGM demonstrated high reproducibility and an LoD of 5% for aneuploidies, translocations, interstitial deletions, and duplications at 400x coverage. OGM identified several additional SVs, revealing the genomic architecture in these neoplasms that can help better classify these neoplasms and predict outcomes. Considering the technical and analytical advantages of OGM compared to the current SOC methods used for chromosomal characterization, we recommend OGM as a potential first-tier cytogenetic test for the evaluation of hematological neoplasms.

## Data Availability

All data produced in the present work are contained in the manuscript and its supplementary files

## Ethics approval

The study was approved by the IRB A-BIOMEDICAL I (IRB REGISTRATION #00000150), Augusta University. HAC IRB # 611298. Based on the IRB approval, the need for consent was waived; all PHI was removed, and all data was anonymized before accessing for the study.

## Declarations

RK has received honoraria, and/or travel funding, and/or research support from Illumina, Asuragen, QIAGEN, Perkin Elmer Inc, Bionano Genomics, Agena, Agendia, PGDx, Thermo Fisher Scientific, Cepheid, and BMS. JH, AH, and AC are salaried employee at Bionano Genomics Inc. All other authors have no competing interests to disclose.

---

**Table 1.** Performance metric evaluation of optical genome mapping for hematological neoplasms.

| Performance Criteria | Overall SVs (n=164) | Aneuploidy (n=61) | Translocation (n=28) | Deletion (n=34) | Duplication/gains (n=11) | Additional material/insertions (n=15) | Marker (n=12) | Isochromosome (n=1) | Ring (n=1) | Inversion (n=1) |
| --- | --- | --- | --- | --- | --- | --- | --- | --- | --- | --- |
| Sensitivity (%) | 98.7 | 98.3 | 100 | 100 | 100 | 93.3 | 100 | 100 | 100 | 100 |
| Specificity (%) | 100 | 100 | 100 | 100 | 100 | 100 | 100 | 100 | 100 | 100 |
| PPV/<br>Precision (%) | 100 | 100 | 100 | 100 | 100 | 100 | 100 | 100 | 100 | 100 |
| NPV (%) | 98 | 100 | 90 | 100 | 100 | 90.9 | 100 | 100 | 100 | 100 |
| Accuracy (%) | 99.2 | 98.5 | 100 | 100 | 100 | 96 | 100 | 100 | 100 | 100 |

**Sensitivity/Positive percentage agreement (PPA) = TP/ (TP+FN).**
**Specificity/Negative percentage agreement (NPA) = TN/ (TN+FP).**

